# Prevalence and Clinical Outcomes of Acute Noncardiac Organ Failure among Non-Acute Myocardial Infarction Cardiogenic Shock: A Nationwide Cohort Analysis

**DOI:** 10.1101/2024.05.20.24307647

**Authors:** Monil Majmundar, Mohammed Faisaluddin, Mohammad Alarfaj, Asmaa Ahmed, Kunal N Patel, Vidit Majmundar, Rajkumar Doshi, Hirak Shah, Kamal Gupta, Zubair Shah, Tarun Dalia

## Abstract

**Background:** Noncardiac organ failure often complicates cardiogenic shock (CS). The results of cardiogenic shock caused by noncardiac organ failures in patients without acute myocardial infarction (AMI) are not well documented.

**Methods:** We examined the National Inpatient Sample (NIS) data from 2016 to 2020 to identify cases of CS and non-AMI CS-related hospitalizations. We divided both cohorts based on the number of acute noncardiac organ failures and evaluated the influence of organ failure on the primary outcome, which was in-hospital mortality.

**Results:** A total of 599,210 (100%) cardiogenic shock and 366,905 (61.2%) non-AMI CS hospitalizations were identified. Among those with non-AMI CS, 58,965 (16.07%) had no organ failure, 121,845 (33.21%) had a single organ failure, and 186,095 (50.72%) had a multiorgan failure. Acute Kidney Injury (AKI) was the most common non-cardiac organ failure (38.1%). Multiorgan failure was associated with an increased risk of in-hospital mortality (aOR: 4.91, 95% CI: 4.72-5.06, p <0.001) compared to no organ failure. A notable increase in mortality rates was observed in correlation with the number of organ involvement. The highest mortality rates were seen in cases where five or more organs were affected. Neurological failure exhibited a significant association with mortality when compared to other organ failures. Similar trends were seen among the CS cohort.

**Conclusions:** In non-acute myocardial infarction and all cardiogenic shock patients, AKI is the most common type of organ failure, and neurological failure was associated with the highest mortality rate. The presence of noncardiac multiorgan failure was found to be strongly associated with a higher mortality rate. This risk increased as more organs were affected.

**Clinical Perspective:** *What Is New?:*

● Non-AMI patients bear a considerable load of non-cardiac organ failure, where AKI is the most prevalent form of organ failure. Additionally, neurological failure has the highest in-hospital mortality rate.
● A predictor model for calculating the probability of in-hospital mortality in all CS patients.
● Non-AMI cardiogenic shock and all cardiogenic shock patients are associated with higher morbidity, mortality, and resource utilization, as well as advanced circulatory support therapies, which increase as the number of affected organs increases.

*What are the Clinical Implications?:*

● Multi-organ failure in non-AMI CS can have worse cardiovascular outcomes.
● A multidisciplinary team’s involvement in managing these complex CS patients should be considered.

## Introduction

Cardiogenic shock (CS) is a state of inadequate tissue perfusion due to cardiac dysfunction, most commonly caused by acute myocardial infarction (AMI)^1^. Non-AMI CS remains a significant cause of CS-related hospitalizations. The spectrum of non-AMI-CS etiologies is vast and includes end-stage cardiomyopathy, myocarditis, valvular heart disease, and pericardial disease^2^. In a study by González-Pacheco et al. among non-AMI CS patients, valvular heart disease (49.3%) and cardiomyopathies (42.3%) constituted primary etiologies for the non-AMI CS. CS is often associated with organ failure, which can directly and indirectly impact the prognosis, including mortality^3^. A study by Vallabhajosyula et al. reported 32.4% and 31.9% prevalence for single-organ and multi-organ failure, respectively, among AMI-CS patients^4^. Furthermore, multi-organ failure was associated with higher in-hospital mortality (48%) and was also associated with increased need for Mechanical circulatory support (MCS) and longer length of hospital stay. Recently, the Detroit Cardiogenic Shock Initiative showed a reduction in in-hospital mortality among CS patients to (24%) on incorporating the CS treatment algorithms, which included early recognition of CS, early revascularization, early use of invasive hemodynamic monitoring, and early deployment of MCS^5^. Despite refinements in CS treatment strategies, mortality among patients with CS continues to be substantial^6^. Heart Failure is one of the potential major contributors of CS in the non-AMI population. The Non-AMI CS differs in characteristics and outcomes from AMI-CS^7^. The data regarding mortality among non-AMI CS patients is variable. The mortality risk is driven by many factors, including shock severity, response to therapy, the magnitude of organ dysfunction, and, more importantly, the type of organ involved^8^. Hence, a comprehensive assessment of shock severity should be done on the factors mentioned above. Therefore, using the nationally representative inpatient database, we sought to determine the burden of organ failure associated with non-AMI CS, the frequency of organ involvement and its association with mortality, and its implications regarding resource utilization compared to the organ failure associated with all CS.

## METHODS

We analyzed data from the National Inpatient Sample (NIS) database from 2016 to 2020. NIS is part of the healthcare cost and utilization project (HCUP) databases. These databases are sponsored by the Agency for Healthcare Research and Quality (AHRQ)^9^. The NIS database represents nearly 95% of the US population and includes 20% of discharge patient data. The NIS undergoes annual quality assessments to confirm its internal validity. Additionally, the NIS is a publicly available database with de-identified data. Therefore, Institutional Review Board approval was not required for our study.

### Study Population and Design

We selected the CS cohort using the International Classification of Disease, Tenth Edition, Clinical Modification (ICD-10-CM) code “R570”. Of the total **849,230** hospitalizations, we excluded patients age <18 years of age (N=11,745), COVID infection (N=10,435), Sepsis/septic shock patients (N=179,905), and end-stage renal disease (ESRD) (N=45,545). Patients with COVID, septic shock, and ESRD were removed from this analysis to avoid any confounders from such diseases. Subsequently, we used ICD-10 variables to define different organ failures, which include neurological failure, respiratory failure, acute kidney injury, hepatic failure, and hematological failure. Definitions of each variable and code are listed in **Table S1**. We subcategorize organ failure into single-organ failure, which consists of any organ failure mentioned above. Multi-organ failure was defined as the presence of two or more organ failures. The comorbidities included are detailed in **Table S2**, encompassing all the comorbidities listed in the Elixhauser Comorbidities Index. The cohort selection flow diagram is shown in **Supplemental Figure S1**.

### Study Outcomes

Cardiovascular outcomes were demonstrated in no-organ failure, single-organ failure, and multi-organ failure, which formed the three groups for each non-AMI and all CS cohorts. The primary outcome measured was in-hospital mortality. Secondary outcomes included cardiac arrest, ventricular tachycardia (VT), total hospitalization cost, length of stay, and disposition. We also studied resource utilization, which included mechanical circulatory support (MCS) use, coronary angiography, Left Ventricular Assist Devices (LVADs), and Heart Transplants (HTs). We also generated a predictor model for in-hospital mortality. The common variable definitions of primary and secondary outcomes with their ICD-10 codes are shown in **Supplemental Table S1-S3**.

### Statistical Analysis

As recommended, survey procedures using discharge weights provided with the HCUP-NIS databases were used to generate national estimates. Descriptive statistics were used to analyze the demographic and comorbidity data. Most hospital-level characteristics were directly obtained as provided in the NIS, whereas the Elixhauser Comorbidity Index was used to identify comorbid disorders. Descriptive statistics were presented as frequencies with percentages for categorical variables and medians with interquartile range (IQR) for continuous variables. Baseline characteristics were compared using Pearson X2 and Fisher’s exact tests for categorical variables. Mann-Whitney U-Test was used for continuous variables presented as the median and interquartile range (IQR). We conducted a multivariable logistic regression using the Cochran-Mantel-Haenszel test to control for baseline demographics, comorbidities, and hospital characteristics to evaluate clinical outcomes during hospitalization between the three groups with the no-organ failure group as the reference. We used a logistic regression model to create a predictor model of in-hospital mortality probability. Detailed variables used for adjusted analysis are presented in **Table S3**. To estimate hospital cost (total expense for hospital services), the NIS-provided “cost-to-charge ratio” variable was multiplied by the total inpatient charges (the total billed amount by the participating hospital) and was adjusted for inflation wages of January 2020 using the Bureau of Labor Statistics Consumer Price Index (adjusted cost) ^10^. A p-value of <0.05 with a 95% confidence interval (CI) not crossing “1” was set as statistical significance for any acceptable error <5% to disprove the null hypothesis. All statistical analyses were performed with SAS 9.4 (SAS Institute, Inc., Cary, North Carolina)

## RESULTS

884,920 CS hospitalizations were identified, of which 599,210 met our inclusion and exclusion criteria. The majority of those, 366,905 (61.2%), were non-AMI CS hospitalizations. Among those with non-AMI CS, a total of 58,965 (16.07%) had no organ failure, 121,845 (33.21%) had a single organ failure, and 186,095 (50.72%) had a multiorgan failure. In the CS cohort, a total of 110,035 (18.36%), 190,535 (31.8%), and 298,640 (49.84%) had no organ failure, single organ failure, and multiple organ failure, respectively.

### Demographic and baseline comorbidities

Among non-AMI CS hospitalizations, the median age for patients with no organ failure was 66 years, while for those with a single organ failure and multiorgan failure, it was 67 years. Caucasians (68.03%) and males (62.1%) constituted most cases. Hypertension, heart failure, diabetes, hyperlipidemia, and atrial fibrillation were common comorbidities in CS and non-AMI CS. Demographic and baseline comorbidities for both cohorts are detailed in **(Table 1)**.

**Table 1:** Baseline characteristics of Non-AMI CS and CS patients stratified by number of organ failure; Abbreviations: - Non-AMI indicates Non-Acute Myocardial Infarction, PAD: Peripheral Arterial Disease, HTN: Hypertension, CKD: Chronic Kidney Disease, CHF: Congestive Heart Failure, COPD: Chronic Obstructive Pulmonary disease.

|  | *Cardiogenic shock (n=599,210, 100%)* |  |  |  | *Non-AMI Cardiogenic Shock (n=366905, 61.2%)* |  |  |  |
| --- | --- | --- | --- | --- | --- | --- | --- | --- |
|  | *No organ failure*<br>*110035*<br>*(18.36%)* | *Single organ failure*<br>*190535*<br>*(31.80%)* | *Multi-organ failure*<br>*298640*<br>*(49.84%)* | *P value* | *No organ failure*<br>*58965*<br>*(16.07%)* | *Single organ failure*<br>*121845*<br>*(33.21%)* | *Multi-organ failure*<br>*186095*<br>*(50.72%)* | *P value* |
| Age (Median) | 66 (57-76) | 68 (58-77) | 68 (58-77) |  | 66 (56-76) | 67 (57-77) | 67 (57-76) |  |
| Female (%) | 38.39 | 37.42 | 35.43 | <.0001 | 39.31 | 37.32 | 36.84 | <.0001 |
| ***Race*** |  |  |  | <.0001 |  |  |  | <.0001 |
| Caucasian (%) | 72.75 | 70.13 | 69.91 |  | 69.10 | 67.04 | 67.95 |  |
| African American (%) | 12.19 | 15.60 | 14.88 |  | 17.17 | 19.60 | 17.79 |  |
| Hispanic (%) | 8.12 | 7.87 | 8.18 |  | 8.07 | 7.53 | 7.85 |  |
| Asian | 3.01 | 2.90 | 3.10 |  | 2.40 | 2.65 | 2.75 |  |
| Native Americans | 0.61 | 0.58 | 0.62 |  | 0.57 | 0.57 | 0.55 |  |
| Others | 3.32 | 2.91 | 3.31 |  | 2.68 | 2.60 | 3.10 |  |
| ***Comorbidities*** |  |  |  |  |  |  |  |  |
| COPD | 22.92 | 28.06 | 27.52 | <.0001 | 26.49 | 30.01 | 29.70 | <.0001 |
| Obesity | 16.73 | 18.76 | 19.04 | <.0001 | 17.98 | 19.73 | 20.15 | <.0001 |
| Hyperlipidemia | 55.64 | 51.46 | 43.02 | <.0001 | 50.03 | 47.45 | 39.12 | <.0001 |
| Alcohol | 4.61 | 5.38 | 6.69 | <.0001 | 5.54 | 5.76 | 7.52 | <.0001 |
| CHF | 65.26 | 77.02 | 76.77 | <.0001 | 74.71 | 80.72 | 77.79 | <.0001 |
| Valvular heart disease | 26.38 | 30.35 | 27.16 | <.0001 | 35.36 | 35.56 | 31.37 | <.0001 |
| Prior Valve Surgery | 2.54 | 2.89 | 2.48 | <.0001 | 3.82 | 3.90 | 3.18 | <.0001 |
| PAD | 18.05 | 19.58 | 16.88 | <.0001 | 22.58 | 21.37 | 17.74 | <.0001 |
| Coagulopathy | 3.09 | 11.99 | 36.32 | <.0001 | 3.84 | 13.28 | 39.78 | <.0001 |
| Liver Disease | 3.11 | 6.45 | 31.72 | <.0001 | 4.19 | 7.73 | 32.11 | <.0001 |
| Pulmonary<br>circulatory<br>disorder | 16.06 | 21.89 | 21.93 | <.0001 | 24.55 | 28.28 | 27.72 | <.0001 |
| Metastatic<br>Cancer | 3.31 | 3.87 | 3.56 | <.0001 | 4.11 | 4.32 | 4.06 | <.0001 |
| Atrial<br>fibrillation | 33.52 | 42.11 | 42.87 | <.0001 | 41.60 | 48.29 | 47.68 | <.0001 |
| Coronary artery<br>disease | 24.71 | 25.46 | 19.7 | <.0001 | 27.85 | 26.55 | 19.65 | <.0001 |
| Conduction<br>Disorders | 15.14 | 16.00 | 15.93 | <.0001 | 13.73 | 15.27 | 14.46 | <.0001 |
| HTN | 71.52 | 77.24 | 74.51 | <.0001 | 72.44 | 77.69 | 74.36 | <.0001 |
| Diabetes | 34.40 | 39.78 | 37.97 | <.0001 | 34.74 | 38.45 | 35.32 | <.0001 |
| Smoking | 24.98 | 24.77 | 18.72 | <.0001 | 27.61 | 26.24 | 19.14 | <.0001 |
| Drug abuse | 4.62 | 5.25 | 6.12 | <.0001 | 5.56 | 5.95 | 7.26 |  |
| CKD | 17.99 | 35.38 | 37.36 | <.0001 | 24.86 | 41.29 | 40.41 | <.0001 |
| Acute<br>Myocardial<br>Infarction | 46.41 | 36.05 | 37.68 | <.0001 | NA | NA | NA | NA |
| **Primary payer** |  |  |  | <.0001 |  |  |  | <.0001 |
| Medicare | 55.73 | 61.55 | 59.73 |  | 57.43 | 60.94 | 58.95 |  |
| Medicaid | 11.44 | 11.46 | 11.88 |  | 13.35 | 12.81 | 13.27 |  |
| Private<br>insurance | 25.37 | 20.76 | 21.39 |  | 23.50 | 20.71 | 21.10 |  |
| other | 2.88 | 2.79 | 2.88 |  | 2.54 | 2.77 | 2.85 |  |
| **Bed-size of the**<br>**hospital** |  |  |  | <.0001 |  |  |  | <.0001 |
| Small | 13.60 | 12.92 | 12.75 |  | 12.31 | 12.15 | 12.37 |  |
| Medium | 24.66 | 24.25 | 24.86 |  | 21.02 | 21.92 | 23.44 |  |
| Large | 61.74 | 62.82 | 62.39 |  | 66.67 | 65.92 | 64.19 |  |
| **Teaching**<br>**status of the**<br>**hospital** |  |  |  | <.0001 |  |  |  | <.0001 |
| Rural hospital | 5.65 | 4.48 | 3.34 |  | 4.94 | 3.73 | 3.10 |  |
| Urban non-<br>teaching | 16.98 | 15.53 | 15.74 |  | 13.81 | 13.38 | 14.48 |  |
| Urban teaching | 77.37 | 79.99 | 80.93 |  | 81.24 | 82.89 | 82.41 |  |
| **Hospital**<br>**Region** |  |  |  |  |  |  |  |  |
| Northeast | 17.66 | 17.17 | 15.53 |  | 18.02 | 17.34 | 15.79 |  |
| Midwest | 22.49 | 23.24 | 23.34 |  | 22.82 | 23.63 | 23.70 |  |
| South | 40.46 | 39.42 | 39.60 |  | 40.77 | 39.09 | 39.14 |  |
| West | 19.39 | 20.18 | 21.53 |  | 18.39 | 19.94 | 21.36 |  |

**Table 2:** Resource Utilization of Non-AMI CS and CS patients stratified by the number of organ failure; Abbreviations: - PCI: Percutaneous Coronary Intervention, CABG: Coronary Artery Bypass Graft, ECMO: Extracorporeal Membrane Oxygenation, IABP: Intra-aortic Balloon Pump, pVAD: Percutaneous Ventricular Assist Device, LVAD: Left Ventricular Assist Device, MCS: Mechanical Circulatory Support.

|  | *Cardiogenic shock (n=599,210, 100%)* |  |  |  | *Non-AMI Cardiogenic Shock (n=366905, 61.2%)* |  |  |  |
| --- | --- | --- | --- | --- | --- | --- | --- | --- |
| Outcomes | *No organ failure*<br>*110035*<br>*(18.36%)* | *Single organ failure*<br>*190535*<br>*(31.80%)* | *Multi-organ failure*<br>*298640*<br>*(49.84%)* | *P value* | *No organ failure*<br>*58965*<br>*(16.07%)* | *Single organ failure*<br>*121845*<br>*(33.21%)* | *Multi-organ failure*<br>*186095*<br>*(50.72%)* | *P value* |
| Angiogram | 46.33 | 34.67 | 32.32 | <.0001 | 20.69 | 17.81 | 16.51 | <.0001 |
| PCI | 30.82 | 18.53 | 17.43 | <.0001 | 4.34 | 2.97 | 2.55 | <.0001 |
| CABG | 13.19 | 11.29 | 9.26 | <.0001 | 11.06 | 8.09 | 6.23 | <.0001 |
| ECMO | 0.33 | 0.61 | 2.15 | <.0001 | 0.31 | 0.62 | 2.10 | <.0001 |
| IABP | 18.05 | 15.05 | 15.69 | <.0001 | 6.85 | 7.18 | 8.19 | <.0001 |
| pVAD | 5.23 | 4.75 | 7.30 | <.0001 | 2.15 | 2.00 | 3.51 | <.0001 |
| MCS | 23.61 | 20.41 | 25.14 | <.0001 | 9.31 | 9.8 | 13.8 | <.0001 |
| LVAD-Implantable | 1.29 | 1.97 | 2.06 | <.0001 | 2.33 | 2.94 | 2.94 | <.0001 |
| Heart Transplant | 0.58 | 1.15 | 1.20 | <.0001 | 1.09 | 1.77 | 1.86 | <.0001 |
| Invasive hemodynamic assessment | 17.17 | 19.14 | 16.19 | <.0001 | 22.51 | 21.54 | 21.54 | <.0001 |

### Hospital resource utilization

In non-AMI-CS, patients with multiorgan failure had the highest rates of MCS support with intra-aortic balloon pump (IABP) (no organ failure: 6.85%, one organ failure: 7.2%, multiorgan failure: 8.2%), extracorporeal membrane oxygenation (ECMO) (no organ failure:0.3%, one organ failure: 0.6%, multiorgan failure: 2.1%), and percutaneously inserted ventricular assist device (pVAD) (no organ failure: 2.2%, one organ failure: 2.0%, multiorgan failure: 3.5%). Increased use of Left Ventricular Assist Devices (LVADs) and Heart Transplants (HTs) (no organ failure: 2.33%, one organ failure: 2.94%, multiorgan failure: 2.94% for LVADs) and no organ failure: 1.09%, one organ failure: 1.77%, multiorgan failure: 1.86% for HTs) were also noted in the Non-AMI-CS population with involvement of organ dysfunction. However, those with multiorgan failure had less frequent coronary procedures, including a coronary angiogram, PCI, and CABG, compared to no organ failure and single organ failure. All resource utilization for both cohorts is detailed in (**Table. 2**)

### In-Hospital Outcomes

Respiratory and renal failures were the most involved systems in all CS (36.5% and 33.2%, respectively) and non-AMI CS patients (38.1% and 30.8%, respectively). Patients with four or five organ involvement, as well as individuals with neurological failure and hepatic failure, experienced the highest mortality rates overall **(Figures 2 A, and 2 B**).

**Figure 1.**
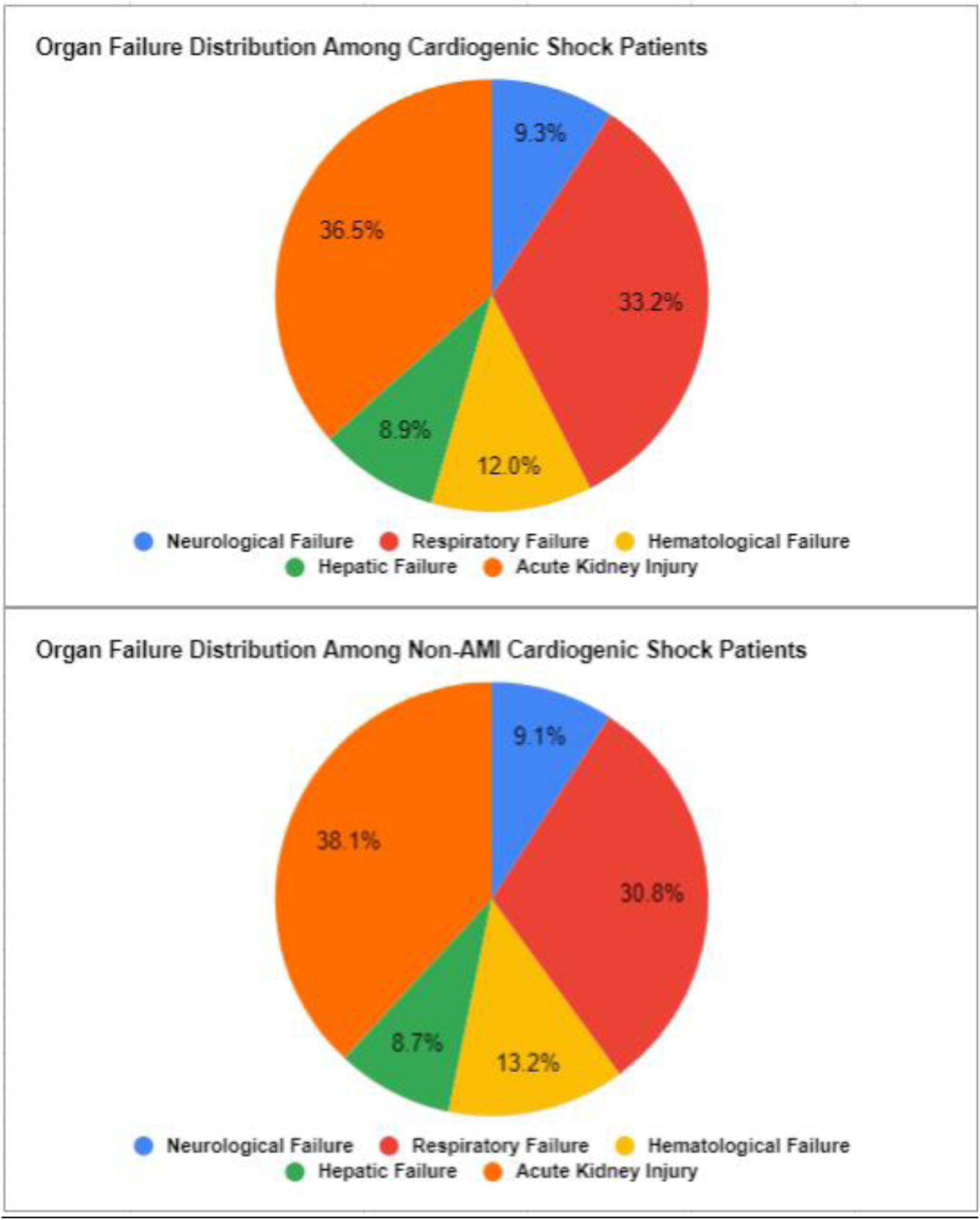
Pie chart showing the distribution of type of organ failure in CS and non-AMI CS. Respiratory failure and Acute Kidney Injury were the most common types of organ failure noted in both subgroups.

**Figure 2.**
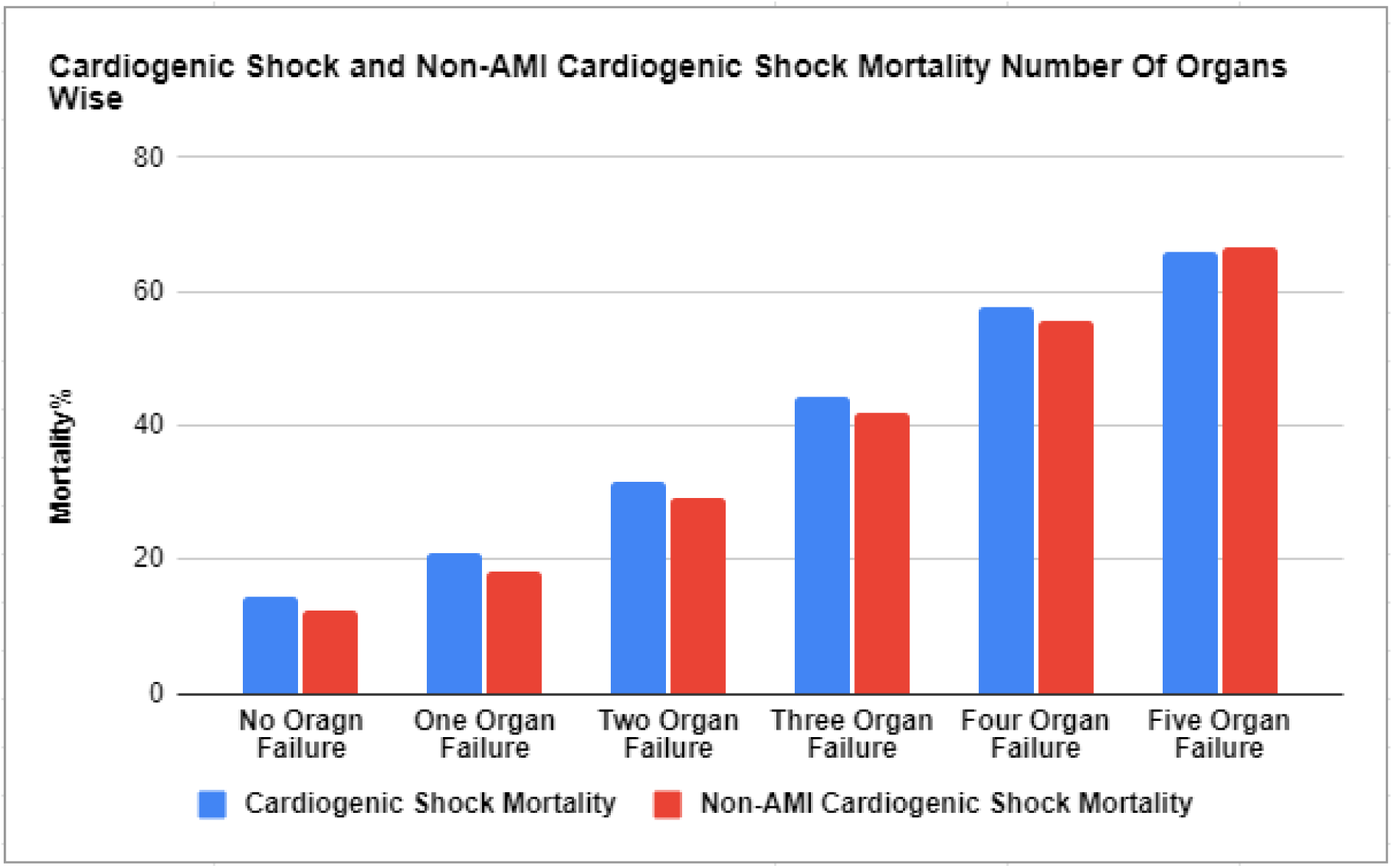
A Bar diagram showing in-hospital mortality by number of organ failures in cardiogenic shock and non-AMI cardiogenic shock patients. Mortality increases proportionately as the number of organ failures increases from no organ failure to five organ failure.

**Figure 2.**
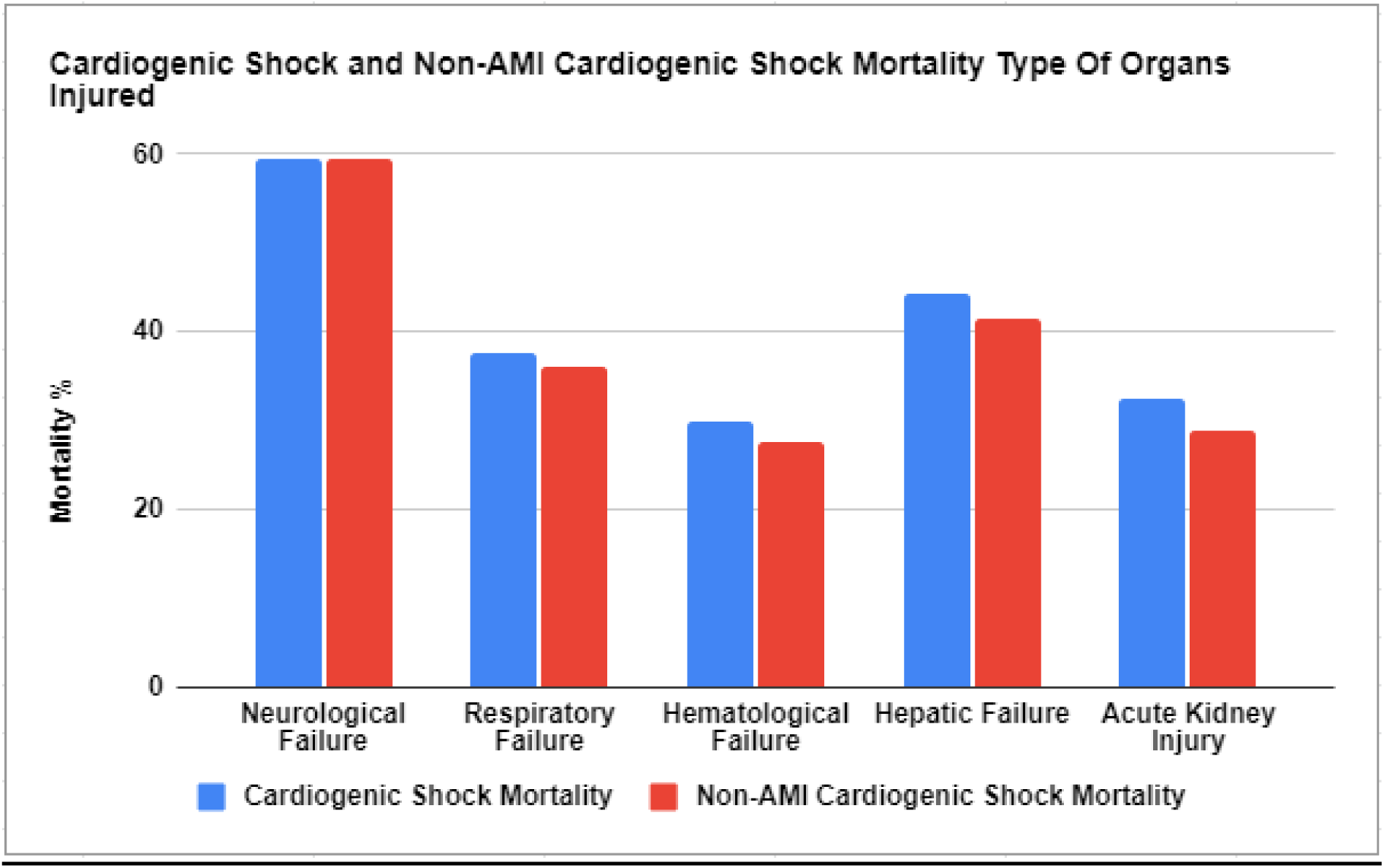
B; Bar diagram showing in-hospital mortality by type of organ failure in cardiogenic shock and non-AMI cardiogenic shock patients. Neurological failure was associated with the highest correlation to mortality in both Non-AMI CS and all CS patients.

In a multivariate analysis, the cohort with single organ failure and multiorgan failure had higher rates of in-hospital mortality (aOR: 1.84; 95% CI: 1.78-1.90, <.0001) and (aOR: 4.91; 95% CI: 4.72-5.06, <.0001), respectively, cardiac arrest (aOR 1.54;1.45-1.63,<.0001) and (aOR 2.90;2.73-3.07,<.0001), respectively. In-hospital outcomes for both cohorts are highlighted in (**Table 3**). We created a predictor model for the probability of in-hospital mortality in all CS patients, where female gender, presence of complications or comorbidities, and Medicaid insurance get a value of 1 in the equation. The area under the curve is 0.73.

**Table 3.**
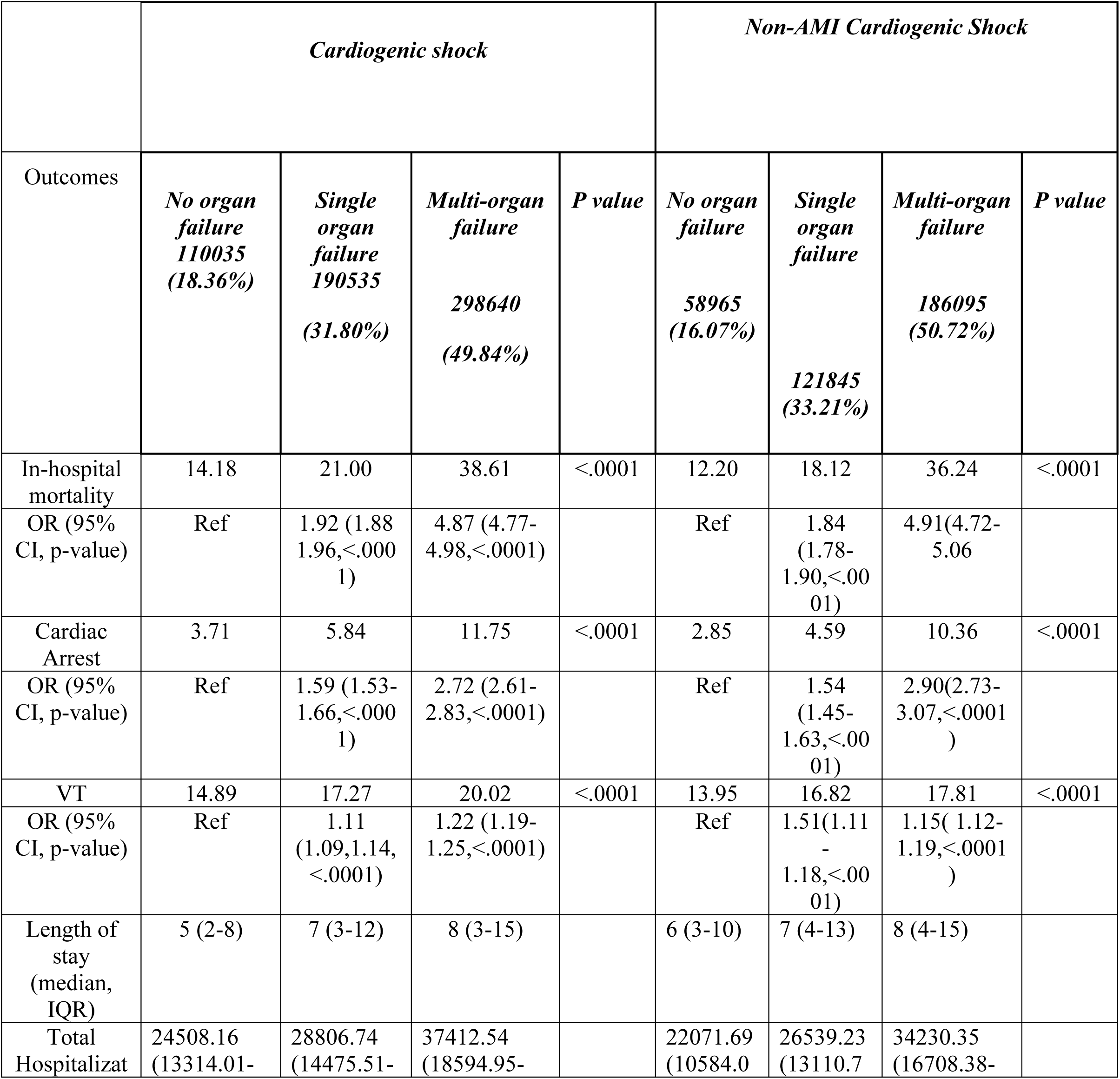
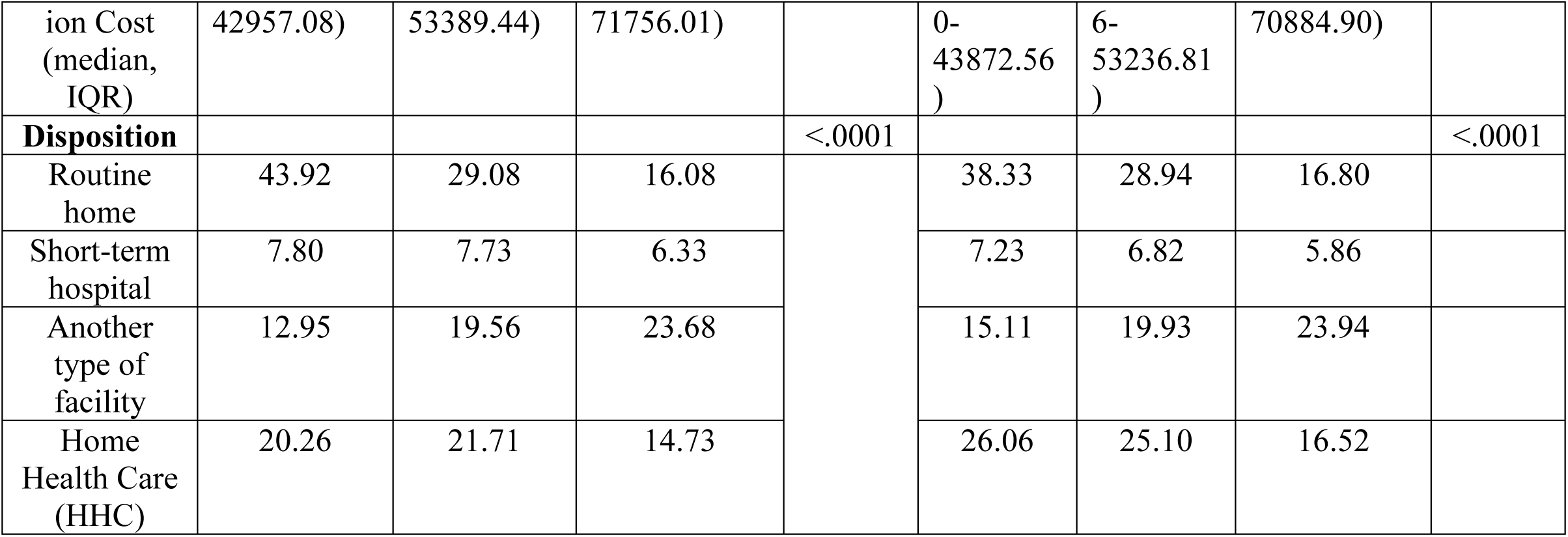
In-hospital Outcomes of Non-AMI CS and CS patients stratified by the number of organ failure

x = -3.75 + 0.03 (Age in years) + 0.20 (Female gender) + 0.08 (Acute MI) + 0.76 (Respiratory failure) + 0.68 (Hepatic failure) + 0.06 (Hematologic failure) + 1.55 (Neurological failure) + 0.37 (AKI) + 0.35 (CHF) + 0.18 (hypertension) + 0.21 (peripheral vascular disease) + 0.12 (Medicaid insurance)

In-hospital mortality probability = e^x^/(1 + e^x^)

### Length of stay, cost of care

In the non-AMI CS cohort, the median length of hospital stay progressively increased from none to single and multiple organ failure (5, 7, and 8 days, respectively). Similarly, the median total hospital cost increased ($22,072, $26,539, and $34,230, respectively). Additionally, the likelihood of being discharged to a nursing facility increased from 15.1% in those without organ failure to 19.9% and 23.9% in those with single and multiple organ failure, respectively. Detailed comparisons of these findings for both cohorts are highlighted in (**Table 3**).

## Discussion

The principal findings of our study are: 1) non-AMI cardiogenic shock constituted 61.2% of all CS cases; 2) Renal and respiratory failure was the most common organ failure types in non-AMI CS (**Figure 3**); 3) Neurological failure was associated with the highest mortality in non-AMI CS; and 4) non-AMI CS patients with multiorgan failure had significantly higher in-hospital mortality, a requirement for temporary MCS (tMCS) support, LVADs, HTs, and increased length of stay.

**Figure 3:**
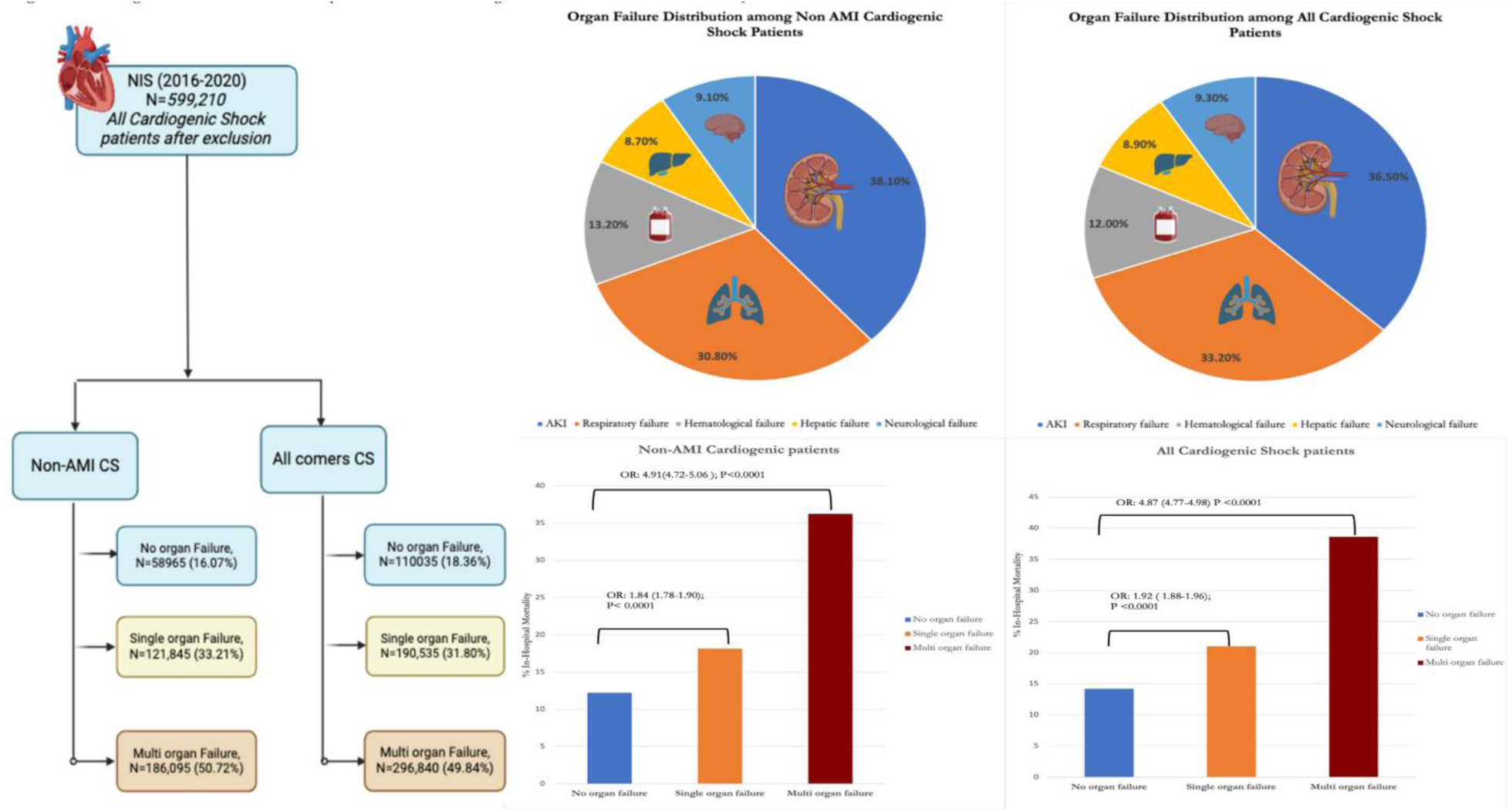
The figure depicts the flow diagram of patient selection along with the most common causes of organ failure among all Cardiogenic Shock and Non-AMI CS patients.

Non-ischemic etiologies remain a significant cause of the reported cases of CS, representing almost 25 to 52% of CS hospitalizations in prior studies^3,11^. Our study found a higher prevalence of non-AMI etiology, accounting for 61.2% of all CS hospitalizations. We found a significant burden of organ failure among non-AMI CS patients. In our study, we found a major burden of chronic heart failure, valvular heart disease, and atrial fibrillation, which is a potential cause for non-AMI cardiogenic shock. Previous studies have documented the relatively similar prevalence of non-AMI CS etiologies^3^.

The spectrum of organ failure in patients with primary cardiac failure is very variable and might range from mild organ dysfunction to more complicated multiorgan failure^12^. In addition, the burden and frequent non-cardiac organ failure in the CS might impact the prognosis, especially in non-AMI CS patients, which is not known. In our study, the most common organ failure was renal failure, followed by respiratory failure and hematological failure.

The impact of noncardiac organ failure in CS has been well documented in the literature and incorporated into the risk stratification and prognosis scores for CS^13,14^. Jentzer et al. validated the relative contribution of each Sequential Organ Failure Assessment (SOFA) organ sub-score for predicting mortality in CICU patients^15^. They reported the cardiovascular, central nervous system, renal, and respiratory SOFA sub-scores to be independently associated with hospital mortality (all p <0.01). In our study, we found neurological failure, followed by hepatic failure and respiratory failure, to be associated with an increased risk of mortality among non-AMI CS patients. Furthermore, with the increasing number of noncardiac organ failures, the risk of in-hospital mortality and cardiac arrest was significantly higher. This was particularly seen with multiorgan involvement, where we found an incremental increase in the mortality risk, with increasing organ involvement highest seen in 5 or more organ failures. Similar findings were reported by Vallabhajosyula et al. among AMI CS patients^4^.

Timely use of early tMCS has been shown to improve outcomes in heart failure-CS patients^7,16^. Approximately 70-80% of the population developing CS in the non-AMI group had the presence of Heart Failure (HF) in their baseline characteristics, making it one of the significant contributors of CS in our non-AMI population. Our study used IABP, ECMO, and pVAD to account for the tMCS. The use of tMCS was ∼20-25% in the all CS group and ∼9-14% in the non-AMI CS, along with an expected finding of incremental increase from no-organ dysfunction to multiorgan dysfunction in both groups.

One of the standard exclusion criteria for LVADs and HTs in patients with advanced heart failure is multi-system organ dysfunction^17^. However, recent studies have shown that LVADs may be performed in patients with severe renal insufficiency without an increase in mortality and morbidity^18^. Similarly, liver cirrhosis is no longer a formal contraindication, and select patients can qualify for LVAD, but ‘irreversible liver damage’ is still considered a contraindication^17^. Interestingly, our study noted that the highest use of LVADs was with multiorgan failure and single organ failure as compared to no organ failure in both the non-AMI CS group and the all-CS group. Similar findings were also noted for HTs. Previous studies using the NIS have also shown higher use of LVADs and HTs in the non-AMI CS population^19^. We believe that these numbers would be underestimated since many patients would have outpatient workups and selection for further advanced heart failure therapies like LVADs and HTs. Finally, we also reported significantly higher overall resource utilization costs among multiorgan failure patients, which can be explained by the fact that multiorgan failure patients had a longer hospital stay, increased risk of short-term outcomes, and higher tMCS requirements^4^.

## Limitations

This study has several important limitations. First, since the NIS is an administrative claims database, it is susceptible to coding and documentation errors. However, estimates, clinical characteristics, and procedural data from the NIS have been extensively validated internally and externally. Second, we were unable to report patient-level data on laboratory results and concomitant medication use, which could have provided valuable insight into diagnosis and prognosis. Third, we could not determine the method of cardiogenic shock diagnosis and its severity, which are crucial factors affecting outcomes. Fourth, the lack of data on the determining hemodynamic data like CO/CI, PCWP, and timing of mechanical circulatory support (MCS) usage limited our ability to determine the true prevalence of non-AMI CS etiology and how it impacts outcomes in each organ failure category. Finally, the NIS data are restricted to hospital stay duration, and we cannot conclude long-term outcomes.

## Conclusion

In conclusion, our comprehensive analysis of non-AMI CS and all CS hospitalizations using the National Inpatient Sample showed crucial insights into the impact of noncardiac organ failure on clinical outcomes and medical resource utilization. Notably, patients with multiorgan failure had the highest mortality rates, with neurological failure demonstrating a strong association with adverse outcomes. Furthermore, individuals with multiorgan failure received increased MCS and had a more extended and more costly hospital stay. More dedicated research in patients with non-AMI-CS is needed to prevent and treat multi-organ failure in these patients to improve outcomes.

## Data Availability

https://hcup-us.ahrq.gov/db/nation/nis/nisdbdocumentation.jsp

## Author Contributions

## Abbreviations

CS: Cardiogenic shock
AKI: Acute Kidney Injury
HF: Heart Failure
AMI: Acute Myocardial Infarction
MCS: Mechanical circulatory support
SCA: Sudden Cardiac Arrest
LOS: Length of Stay
DRG: Diagnosis Related Groups
IQR: Interquartile ranges
Elixhauser Comorbidity Index: ECI
HCUP: Healthcare Cost and Utilization Project
NIS: Nationwide Inpatient Sample

---

**Supplemental Figure S1:**
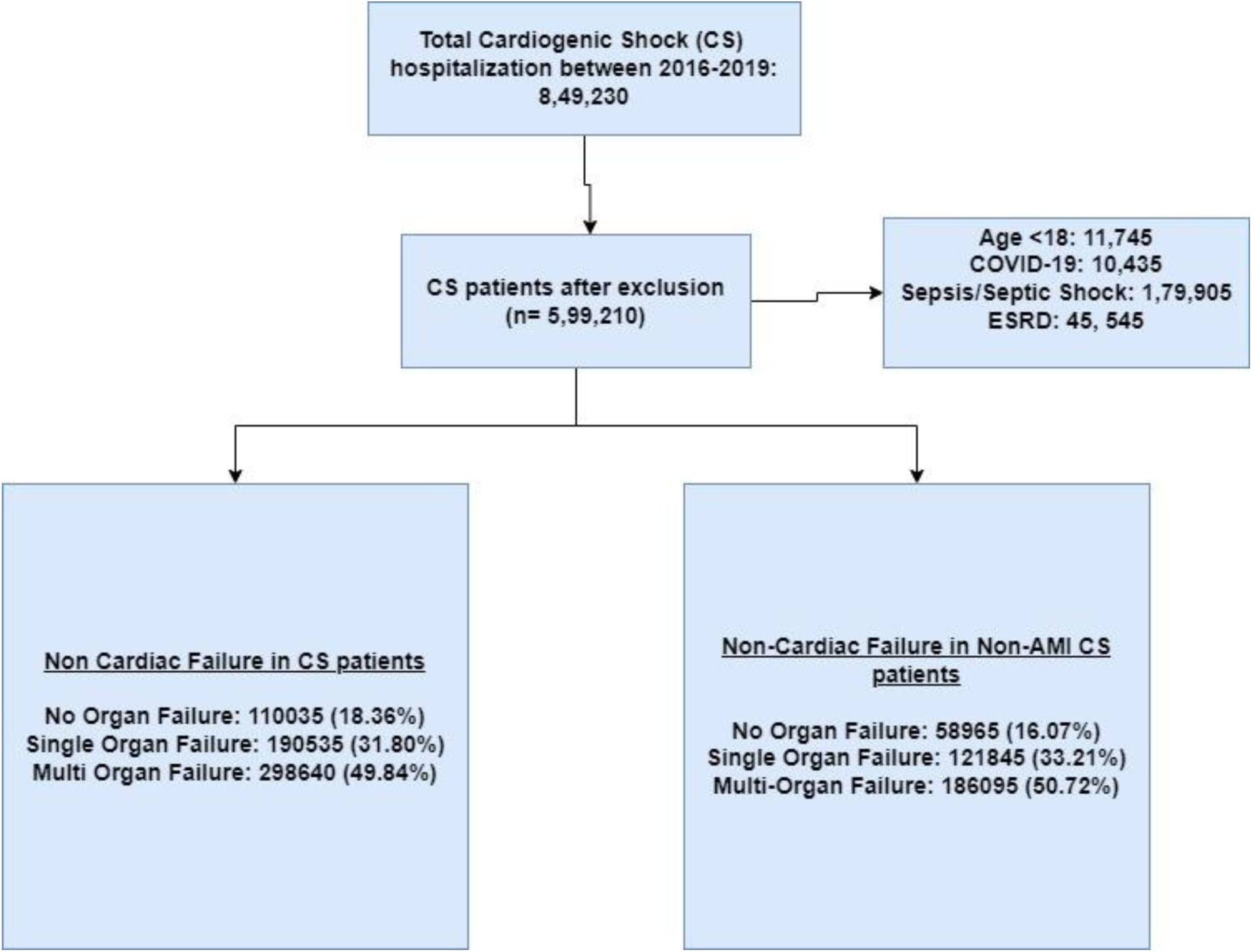
Cohort Flow Selection Diagram

